# Reduced Systemic Homeostatic and Endothelial Protective Capacity in Adolescent Major Depression: Associations with Early-Life Adversity and Clinical Phenotypes

**DOI:** 10.64898/2026.10.11.26365268

**Authors:** Abbas F. Almulla, Ping Wang, Huajie Mao, Yaoyin Zhang, Juan Li, Junmin Chen, Yulan Huang, Yingqian Zhang, Michael Maes

## Abstract

**Background:** Major depressive disorder (MDD) is a heterogeneous condition characterized by neuro-immune-metabolic-oxidative stress (NIMETOX) pathways. A systems-biology approach may help elucidate how these interconnected NIMETOX disturbances relate to early-life adversity and multidimensional clinical phenomes in adolescent MDD.

**Objectives:** This study aimed to define the peripheral metabolic, vascular, and acute-phase, oxidative, and lipid biomarker profiles of adolescent MDD and to investigate associations with ACEs and clinical phenotypes.

**Methods:** This cross-sectional study recruited 80 adolescents with MDD and 53 matched healthy controls and assessed C-peptide and insulin, vascular tone markers, the acute-phase inflammatory axis, oxidative stress, and lipid profiles.

**Results:** MDD adolescents exhibited significant reductions in C-peptide, NO, ApoE, ApoA1, HDL-C, albumin, and transferrin, together with increased ApoB/ApoA1 ratio. These biomarkers correlated significantly with family dysfunction, childhood trauma, bullying exposure, and maladaptive parenting. These biomarkers and stressors accounted for 46.2% to 61.2% of the variance in psychological resilience, and major affective, physiosomatic, self-injurious, suicidal, and self-regulation phenomes. ACEs and bullying exposure, together with relevant socio-demographic data, explained 10.5% to 28.4% of the variance in selected metabolic, vascular, and acute-phase biomarkers. Multivariable classification models showed adequate discrimination between MDD and healthy controls, achieving up to 88.5% classification accuracy and an AUC of 0.948.

**Conclusion:** Adolescent MDD is characterized by coordinated reductions in negative acute-phase proteins, metabolic and lipoprotein protective factors, and endothelial NO, without overt CRP-defined inflammation or conventional dyslipidemia. These findings support a systems-biology model of impaired peripheral homeostatic protection associated with developmental adversity and multidimensional depressive phenomes.

## Introduction

Major depressive disorder (MDD) is not a singular biological phenomenon, and binary diagnostic classification captures only a fraction of the variability in its clinical expression and pathophysiology (Maes, Almulla et al. 2025, Maes, Almulla et al. 2026). Rather than a unitary construct, adolescent depression comprises a spectrum of clinical phenomes—spanning pure affective distress, somatic-vegetative burden, behavioral dysregulation (self-harm, suicidality, and impaired self-control), and compromised psychological resilience (Wang, Wang et al. 2026). Despite the frequent occurrence of these domains amongst developmental stress, the peripheral biological mechanisms connecting early adversity to these clinical traits in adolescents are still inadequately comprehended (Klein, Arnow et al. 2009, Almulla, Algon et al. 2024, Almulla and Maes 2025).

For two decades, the dominant biological explanation of MDD has centered on immune system namely an upregulated immune-inflammatory response system (IRS), typified by M1 macrophage and T helper (Th)-1/Th-17 cytokine signaling, set against a compensatory immunoregulatory system (CIRS) that is often insufficient (Maes and Carvalho 2018, Maes, Almulla et al. 2025). The negative acute-phase inflammatory response in MDD was first described thirty years ago and recently revisited with precision-phenotyping methods, comprising reductions in serum albumin and transferrin that may occur independently from increased C-reactive protein (CRP) (Maes 1993, Almulla, Niu et al. 2025, Chen, Niu et al. 2025, Maes, Niu et al. 2025). Maes et al demonstrated that negative acute-phase proteins (transferrin and albumin), rather than CRP, distinguished MDD from healthy controls (Maes, Niu et al. 2025).

This may reframe the inflammatory component of MDD biology: reductions in negative acute-phase proteins may occur independently of the classical CRP-dominated positive acute-phase response, indicating that these two components of the acute-phase system can be dissociated in MDD. This asymmetry fits within the neuroimmune-metabolic-oxidative (NIMETOX) framework, in which immune-inflammatory, metabolic, and oxidative-nitrosative pathways constitute distinct but interacting components of MDD pathophysiology rather than a unitary inflammatory cascade (Maes, Almulla et al. 2025, Maes, Jirakran et al. 2025, Maes, Almulla et al. 2026).

Accumulating studies indicate that NIMETOX pathway alterations significantly discriminate patients with MDD from healthy controls and explain substantial variance in key MDD phenomes, including overall severity of depression, physiosomatic symptoms, suicidal behaviors, and recurrence of illness (Maes, Almulla et al. 2025, Maes, Jirakran et al. 2025, Niu, Zhang et al. 2025, Maes, Almulla et al. 2026).

MDD has been linked to impaired reverse cholesterol transport (RCT), increased atherogenicity, and heightened protein oxidation (Almulla, Thipakorn et al. 2023, Maes, Jirakran et al. 2025). RCT depends on a small set of apolipoproteins—principally ApoA1 and ApoE—whose depletion is sufficient to shift the ApoB/ApoA1 ratio toward an atherogenic profile even when conventional lipid measures such as triglycerides, LDL-cholesterol, and ApoB itself remain unremarkable (Almulla, Thipakorn et al. 2023, Chen, Niu et al. 2026).

ApoA1 also exerts important vascular effects by stimulating endothelial nitric oxide synthase (eNOS) and NO production; consequently, reduced ApoA1 may contribute to diminished NO bioavailability, a hallmark of early functional endothelial dysfunction (Wang, Mendelsohn et al. 2014, Cabou, Honorato et al. 2017). A simultaneous decrease in ApoA1, ApoE, and NO could therefore compromise endothelium-dependent vasodilation and vascular homeostasis, potentially without activation of other endothelial vasoactive pathways, including the endothelin-1 (ET-1) system (Cellini, Strobbe et al. 2012). This vascular susceptibility has not, to our knowledge, been investigated as a unified characteristic in adolescent MDD, though Tonhajzerova et al. have shown adolescent MDD to be associated with cardiovascular dysregulation and early vascular dysfunction as preclinical markers of elevated cardiovascular risk (Tonhajzerova, Visnovcova et al. 2022).

C-peptide may contribute independently to this vascular impairment. Beyond its role as a co-secreted marker of insulin output, C-peptide stimulates eNOS activity and enhances nitric oxide release from vascular endothelium (Kitamura, Kimura et al. 2003, Wallerath, Kunt et al. 2003). Furthermore, C-peptide limits leukocyte adhesion to the endothelial wall under endothelial stress, an antiatherogenic action distinct from its secretory origin (Scalia, Coyle et al. 2000). Thus, reduced C-peptide may represent a biologically distinct pathway—complementing the ApoA1-linked mechanism—through which NO bioavailability and vascular protection could be compromised (Kitamura, Kimura et al. 2003, Wallerath, Kunt et al. 2003). With its prolonged half-life and minimal first-pass hepatic clearance, C-peptide is a reliable indicator of endogenous secretory activity, and its metabolic and vascular significance in an adolescent population remains unexplored. Within the NIMETOX framework, evaluating C-peptide alongside apolipoproteins and negative acute-phase reactants tests whether adolescent depression involves an early failure of protective metabolic-vascular defenses.

We therefore assessed C-peptide, insulin and glucose; NO and ET-1; lipid and apolipoprotein profiles (triglycerides, total cholesterol, HDL-C, LDL-C, ApoB, ApoE, and ApoA1); acute-phase proteins (albumin, transferrin, and hsCRP); and oxidative stress markers (oxidized HDL and AOPP). We examined whether these biomarkers discriminate MDD from healthy controls, explain key MDD phenomes, and are associated with early-life adversity. We hypothesized that adolescent MDD is characterized by altered endothelial signaling, reduced C-peptide and negative acute-phase proteins, and an unfavorable apolipoprotein profile, forming a peripheral immunometabolic and vascular phenotype associated with developmental adversity and multidimensional clinical symptoms.

## Methods

### Participants

The current case-control, cross-sectional study was conducted at the Sichuan Provincial Center for Mental Health between December 2024 and January 2026. The study protocol was reviewed and approved by the Ethics Committee of the University of Electronic Science and Technology of China (approval no. 31682); written informed consent was obtained from a parent or legal guardian for participants below 18 years of age, together with the adolescent’s own written assent, while participants aged 18 years gave consent independently.

In this study we included 133 participants including 80 patients with MDD and 53 matched healthy controls. Adolescents meeting DSM-5 criteria for MDD, diagnosed by a senior psychiatrist, and a score exceeding 17 on the 24-item Hamilton Depression Rating Scale (HAMD-24) (Hamilton, 1960) were enrolled from the inpatient clinic. Healthy controls were physically and psychiatrically healthy adolescents, matched to the patient group on age, sex, BMI, and education. All participants were required to be 12–18 years old, from both sexes, and right-handed. Individuals with a neurological disorder, a history of significant head trauma, a major medical illness such as diabetes mellitus or an active infectious disease, a second psychiatric diagnosis (including bipolar disorder, a schizophrenia-spectrum condition, autism spectrum disorder, intellectual disability, conduct disorder, or obsessive–compulsive disorder), or a lifetime history of substance misuse other than nicotine were not enrolled. Healthy control groups underwent an equivalent screening process and were excluded for any current or past DSM-5 diagnosis, neurological illness, or serious medical condition.

### Clinical assessments

DSM-5 criteria for MDD were used to define diagnoses and symptom evaluations by a senior child and adolescent psychiatrists using the semistructured interview according to DSM-5. Depressive and anxiety symptoms were assessed using the 24-item Hamilton Depression Rating Scale (HAMD-24) (Hamilton 1960), Children’s Depression Inventory (CDI) (Kovacs 1992), 14-item Hamilton Anxiety Rating Scale (HAMA-14) (Hamilton 1959), and Screen for Child Anxiety Related Emotional Disorders (SCARED) (Birmaher, Khetarpal et al. 1997). Psychological resilience was determined via the Connor-Davidson Resilience Scale (CD-RISC) (Connor and Davidson 2003). Somatic symptoms were additionally assessed using the Patient Health Questionnaire-15 (PHQ-15).

Childhood maltreatment was assessed using the Childhood Trauma Questionnaire–Short Form (CTQ-SF) (Bernstein, Stein et al. 2003), family functioning and parental rearing practices were evaluated using the Family Assessment Device (FAD) (Epstein, Baldwin et al. 1983). Parenting and peer victimization were assessed using the Parent–Child Relationship Diagnostic Test (PCRT) (Zhou and Z. Fang 1993), and Delaware Bullying Victimization Scale, respectively. Suicidal ideation, non-suicidal self-injury, and self-regulation difficulties were assessed using the Beck Scale for Suicide Ideation (BSSI) (Beck, Kovacs et al. 1979), Columbia–Suicide Severity Rating Scale (C-SSRS) (Posner, Brown et al. 2011), Ottawa Self-Injury Inventory (OSI) (Posner, Brown et al. 2011), and Swanson, Nolan, and Pelham Rating Scale-IV (SNAP-IV) (Swanson 1992).

To capture the multidimensional clinical presentation of adolescent depression and developmental stress, standardized phenotypic domain indices were computed following our validated structural model (Wang, Wang et al. 2026). The clinical psychopathology phenome comprised six standardized domain scores: the Pure Affective index was computed by averaging the standardized item scores isolating core depressed mood, guilt, worthlessness, and anhedonia across affective rating scales; the Physiosomatic index was derived from the mean of standardized scores reflecting neurovegetative and somatic burden, including insomnia, fatigue, and autonomic complaints. The NSSI Spectrum index, Integrated Suicidal Ideation Severity index, Self-Regulation Behaviour index, and Resilience index were PCA-derived component scores based on prespecified indicators from the OSI, BSSI/C-SSRS, SNAP-IV, and CD-RISC, respectively (Wang, Wang et al. 2026). Similarly, developmental stress exposures were operationalized into four distinct standardized composite scores namely the Family Dysfunction index, the Maladaptive Control Parenting index, the Bullying Exposure index, and the Childhood Trauma index were derived from the corresponding scale or subscale indicators using PCA (Wang, Wang et al. 2026).

### Blood Sampling and Biomarker Assays

Each participant provided a fasting venous blood sample on the morning of clinical assessment, drawn between 07:00 and 09:00 am. Blood was collected into serum-plasma tubes and, after clotting for 10 minutes at room temperature where applicable, centrifuged at 1,300 ×g for 10 minutes at 4 °C within one hour of collection; serum and plasma aliquots were then stored at −80 °C pending batch analysis, avoiding repeated freeze–thaw cycles. Laboratory staff performing the assays were blinded to each participant’s diagnostic status throughout.

A comprehensive panel of acute-phase proteins, lipid and apolipoprotein parameters, metabolic hormones, and oxidative/nitrosative stress markers was quantified in all participants. Albumin, transferrin, high-sensitivity C-reactive protein (hsCRP), standard lipids (TG, TC, HDL-C, LDL-C), and apolipoproteins (ApoA1, ApoB, ApoE) were measured using automated clinical chemistry analyzers. Nitric oxide (NO), advanced oxidation protein products (AOPP), oxidized HDL (oxHDL), endothelin-1 (ET-1), C-peptide, and insulin were determined using colorimetric assays and enzyme-linked immunosorbent assays (ELISA). The ApoB/ApoA1 ratio and the Atherogenic Index of Plasma (AIP = log10 [TG/HDL-C]) were calculated accordingly. Full technical specifications for all assays—including reagent manufacturers, analytical instruments, sensitivities, and intra- and inter-assay coefficients of variation—are detailed in Electronic Supplementary File (ESF) Table 1. In addition, NAPR composite was computed by sum of the standardized z-scores of serum albumin and serum transferrin (zAlbumin + zTransferrin).

**Table 1.**
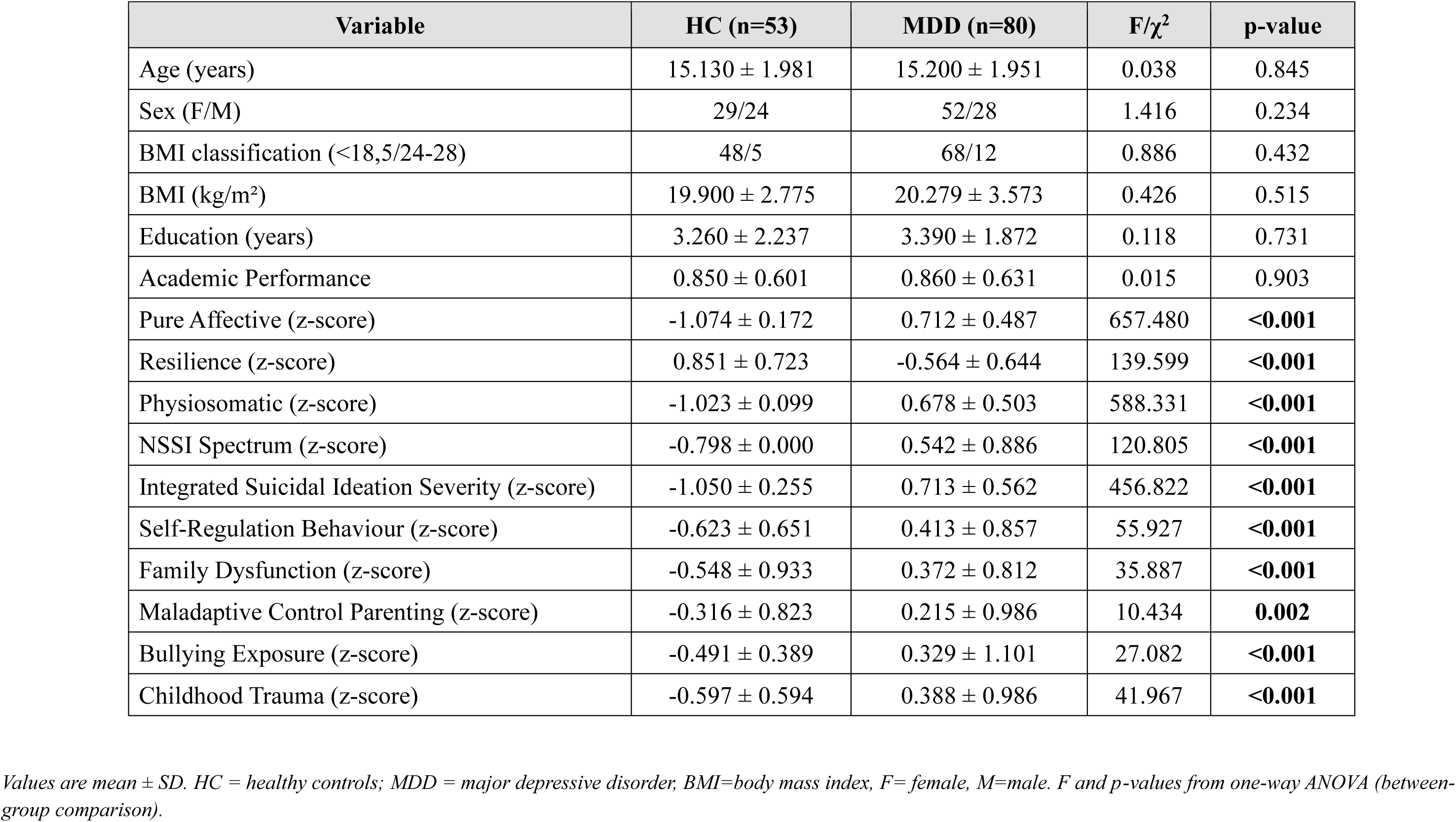
Clinical, socio-demographic, psychosocial, and family characteristics data of adolescents with major depressive disorder (MDD) and healthy controls (HC).

### Statistical Analysis

In this study, we employed IBM SPSS Statistics for Windows, version 30 to conduct all the analyses. Continuous variables were summarized as means and standard deviations or, where comparisons were covariate-adjusted, as estimated marginal means with standard errors; variable distributions were screened beforehand and, where indicated, transformed (log10, square-root, rank-order, or Winsorised) to satisfy model assumptions.

Between-group comparisons treated biomarkers as continuous outcomes within general linear models adjusted for age, sex, and BMI (type III sums of squares) and categorical variables within chi-square tests, with Fisher’s exact test substituted where expected cell counts were small; given comparable group variances and no evidence of meaningful group-by-covariate interaction, the unequal sizes of the MDD and control groups were not treated as a modelling concern. Variance homogeneity was verified from standardised residual plots, group-wise residual distributions, and the Brown–Forsythe test, with linear mixed models (relaxing the equal-variance assumption) applied as a cross-check wherever heteroscedasticity was indicated; resulting p-values were adjusted for the number of biomarkers tested using a false discovery rate (FDR) procedure. Pearson correlation coefficients were used to relate biomarker concentrations to psychosocial, clinical, and demographic variables.

Predictors of individual biomarkers and of the clinical phenotype scores derived in our previous analysis (Wang, Wang et al. 2026) were identified through multiple regression analysis. Multiple regression analyses were performed using demographic, psychosocial, and biomarker variables as candidate explanatory variables. Predictors were selected based on a combination of manual and stepwise regression (entry/removal criteria: p = 0.05/0.10), SPSS Automatic Linear Modeling with overfitting prevention and best-subset selection, and a priori systems-biology knowledge of the pathways under investigation. The selected predictors were subsequently entered simultaneously into a final forced-entry multiple regression model. Each final model is reported with standardized β coefficients, degrees of freedom, R², F, and p values. Coefficient stability was evaluated using 1,000 bootstrap resamples, heteroscedasticity using the White and modified Breusch–Pagan tests, and multicollinearity using tolerance and variance inflation factors.

The capacity of individual and combined biomarkers to discriminate MDD from HC status was assessed by binary logistic regression, with group membership as the dependent variable (HC as reference) and candidate predictors entered stepwise using the likelihood-ratio criterion. Each step is reported with the omnibus model chi-square and Nagelkerke pseudo-R², and each retained predictor with its unstandardised coefficient (B), standard error, Wald statistic, odds ratio (Exp[B]), and 95% confidence interval; classification accuracy is reported at the default 0.5 predicted-probability cutoff. Coefficient stability for the final model was assessed via 1,000-sample bootstrap resampling, yielding bias, standard error, and 95% confidence intervals for each coefficient. Discriminative performance was further quantified from the model’s predicted probabilities using receiver operating characteristic (ROC) curve analysis, reporting the area under the curve (AUC) with its standard error, asymptotic significance, and 95% confidence interval, together with the Gini index; the optimal probability cutoff was identified as the value maximising the Kolmogorov–Smirnov statistic, equivalent to the maximum value of Youden’s index. In parallel, the same discriminative question was addressed by discriminant function analysis, comparing a stepwise solution (Wilks’ lambda entry criterion) against a directly entered model using the full candidate set; for each, standardised canonical coefficients, the structure matrix (pooled within-groups correlations), and group centroids are reported alongside classification accuracy in the original sample and under leave-one-out cross-validation, with coefficient bias, standard errors, and 95% confidence intervals obtained from 1,000-sample bootstrap resampling. Significance for all tests was set at a two-tailed p < 0.05.

## Results

### Sociodemographic, psychological, and clinical characteristics

**Table 1** presents the sociodemographic, clinical, behavioral, and psychological profiles across the study groups. No significant differences were observed between patients with MDD and healthy controls regarding age, sex, BMI, education level, or academic performance.

To evaluate overall differences in clinical presentation while accounting for metabolic status, a two-way multivariate analysis of variance (MANOVA) was conducted on the six standardized clinical psychopathology phenomes, with diagnostic status (MDD vs. healthy controls) and overweight status (BMI-for-age > +1 SD)entered as fixed factors. The analysis revealed a robust, significant multivariate main effect of diagnostic group (Pillai’s Trace = 0.770, F[6, 121] = 67.33, p < 0.001). Neither the main effect of overweight status (Pillai’s Trace = 0.073, F[6, 121] = 1.58, p = 0.158) nor the diagnosis x overweight interaction (Pillai’s Trace = 0.029, F[6, 121] = 0.60, p = 0.727) reached statistical significance.

Subsequent univariate analyses confirmed that adolescent patients with MDD displayed significant elevations across pure affective symptoms, physiosomatic burden, NSSI spectrum behaviors, integrated suicidal ideation severity, and self-regulation behavioral deficits, alongside a marked reduction in psychological resilience. Regarding stress exposures, MDD patients exhibited significantly elevated early-life adversity, characterized by higher levels of family dysfunction, maladaptive control parenting, bullying exposure, and childhood trauma.

### Biomarker alterations in adolescent MDD versus healthy controls

**Table 2** displays differences in the measured biomarkers between MDD patients and healthy controls. The results show significant reductions in negative acute-phase reactions indices including serum albumin (F = 87.43, pFDR < 0.001), transferrin (F = 31.80, pFDR < 0.001), and NAPR composite (F = 70.48, pFDR < 0.001), in MDD versus healthy controls. In addition, significantly lowered levels of NO (F = 39.84, pFDR < 0.001), C-peptide (F = 21.08, pFDR < 0.001), HDL-C (F = 15.46, pFDR < 0.001), ApoE (F = 17.18, pFDR < 0.001), and ApoA1 (F = 20.99, pFDR < 0.001) along with elevated ApoB/ApoA1 ratio (F = 7.27, pFDR = 0.017) were observed in MDD patients compared to healthy controls. Effect sizes for the significantly altered biomarkers ranged from 0.05 to 0.41, with the largest observed, in descending order, for albumin, the NAPR composite, NO, transferrin, and C-peptide. Moreover, hsCRP, AIP, TG, TC, LDL-C, ApoB, AOPP, oxHDL, Insulin, glucose, and ET-1 did not significantly change between study groups. All the results were adjusted for age, sex, and BMI.

**Table 2.**
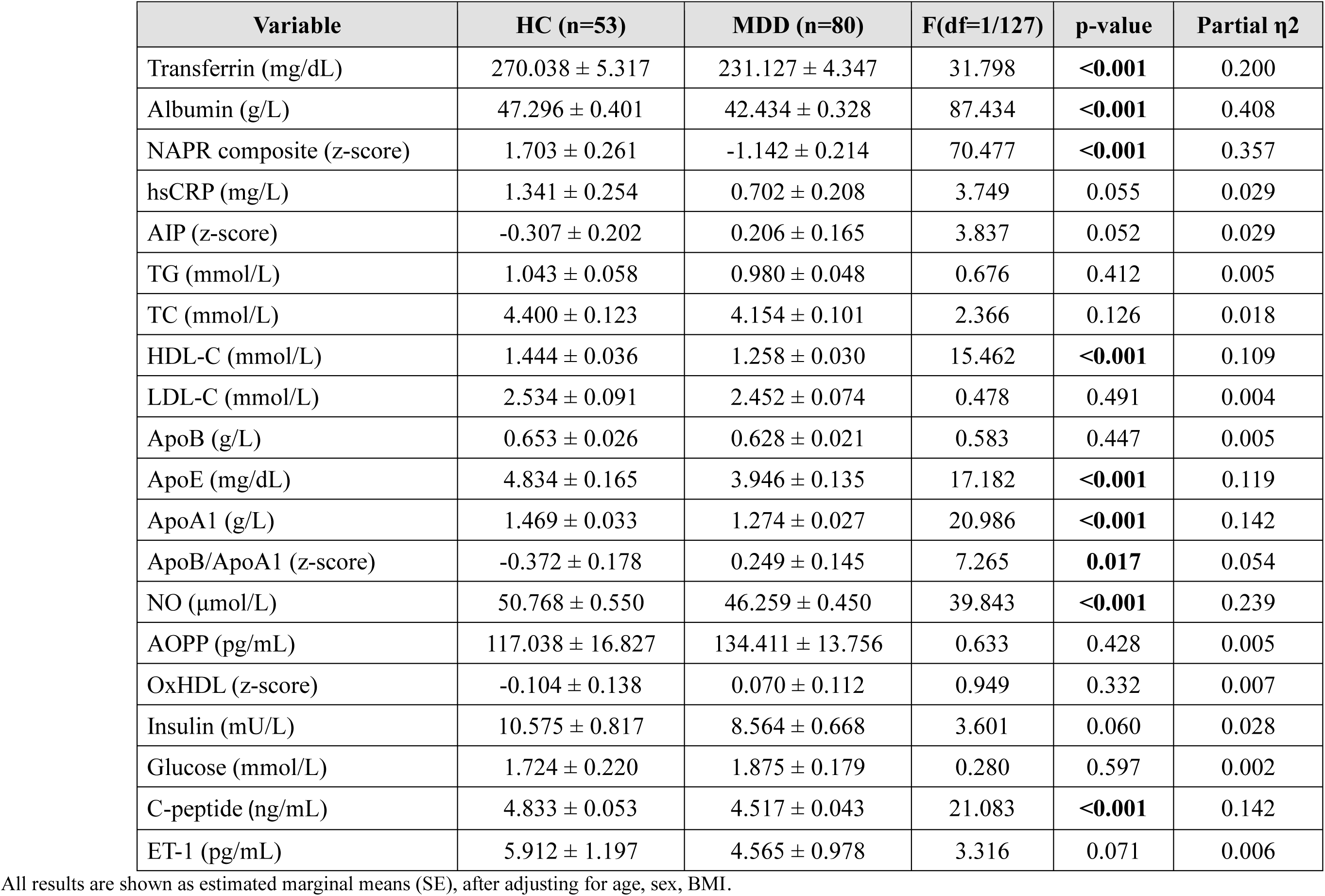
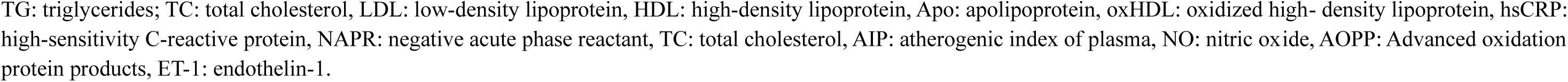
Differences in biomarker profiles between adolescents with major depressive disorder (MDD) and healthy controls (HC). (Estimated Marginal Means)

| Variable | HC (n=53) | MDD (n=80) | F(df=1/127) | p-value | Partial $\eta^2$ |
| --- | --- | --- | --- | --- | --- |
| Transferrin (mg/dL) | 270.038 ± 5.317 | 231.127 ± 4.347 | 31.798 | **<0.001** | 0.200 |
| Albumin (g/L) | 47.296 ± 0.401 | 42.434 ± 0.328 | 87.434 | **<0.001** | 0.408 |
| NAPR composite (z-score) | 1.703 ± 0.261 | -1.142 ± 0.214 | 70.477 | **<0.001** | 0.357 |
| hsCRP (mg/L) | 1.341 ± 0.254 | 0.702 ± 0.208 | 3.749 | 0.055 | 0.029 |
| AIP (z-score) | -0.307 ± 0.202 | 0.206 ± 0.165 | 3.837 | 0.052 | 0.029 |
| TG (mmol/L) | 1.043 ± 0.058 | 0.980 ± 0.048 | 0.676 | 0.412 | 0.005 |
| TC (mmol/L) | 4.400 ± 0.123 | 4.154 ± 0.101 | 2.366 | 0.126 | 0.018 |
| HDL-C (mmol/L) | 1.444 ± 0.036 | 1.258 ± 0.030 | 15.462 | **<0.001** | 0.109 |
| LDL-C (mmol/L) | 2.534 ± 0.091 | 2.452 ± 0.074 | 0.478 | 0.491 | 0.004 |
| ApoB (g/L) | 0.653 ± 0.026 | 0.628 ± 0.021 | 0.583 | 0.447 | 0.005 |
| ApoE (mg/dL) | 4.834 ± 0.165 | 3.946 ± 0.135 | 17.182 | **<0.001** | 0.119 |
| ApoA1 (g/L) | 1.469 ± 0.033 | 1.274 ± 0.027 | 20.986 | **<0.001** | 0.142 |
| ApoB/ApoA1 (z-score) | -0.372 ± 0.178 | 0.249 ± 0.145 | 7.265 | **0.017** | 0.054 |
| NO (μmol/L) | 50.768 ± 0.550 | 46.259 ± 0.450 | 39.843 | **<0.001** | 0.239 |
| AOPP (pg/mL) | 117.038 ± 16.827 | 134.411 ± 13.756 | 0.633 | 0.428 | 0.005 |
| OxHDL (z-score) | -0.104 ± 0.138 | 0.070 ± 0.112 | 0.949 | 0.332 | 0.007 |
| Insulin (mU/L) | 10.575 ± 0.817 | 8.564 ± 0.668 | 3.601 | 0.060 | 0.028 |
| Glucose (mmol/L) | 1.724 ± 0.220 | 1.875 ± 0.179 | 0.280 | 0.597 | 0.002 |
| C-peptide (ng/mL) | 4.833 ± 0.053 | 4.517 ± 0.043 | 21.083 | **<0.001** | 0.142 |
| ET-1 (pg/mL) | 5.912 ± 1.197 | 4.565 ± 0.978 | 3.316 | 0.071 | 0.006 |

All results are shown as estimated marginal means (SE), after adjusting for age, sex, BMI.
TG: triglycerides; TC: total cholesterol, LDL: low-density lipoprotein, HDL: high-density lipoprotein, Apo: apolipoprotein, oxHDL: oxidized high-density lipoprotein, hsCRP: high-sensitivity C-reactive protein, NAPR: negative acute phase reactant, TC: total cholesterol, AIP: atherogenic index of plasma, NO: nitric oxide, AOPP: Advanced oxidation protein products, ET-1: endothelin-1.

### Diagnostic performance of measured biomarkers

We employed binary logistic regression analysis to evaluate the diagnostic performance of the significant measured biomarkers in discriminating MDD patients from healthy controls (**Table 3)**. MDD was significantly associated with lowered albumin, ApoE and C-peptide as shown in model #1 (χ² = 78.603, df = 3, p < 0.001, Nagelkerke R² = 0.606). The current model demonstrated a high overall classification accuracy (82.6%), sensitivity (87.3%), and specificity (75.5%). ROC analysis showed that this model achieved an AUC of 0.907, with an overall quality of 0.86, and a Youden’s index of 0.659. Furthermore, another model (model #2) included adding hsCRP, ET-1, and family dysfunction improved the overall predictive performance and showed that MDD was associated with reduced biomarkers (albumin, ApoE, C-peptide, hsCRP, ET-1) and increased family dysfunctions (χ² = 101.41, df = 6, p < 0.001, Nagelkerke R² = 0.731) even after 1000 bootstrap. This model showed higher overall discrimination accuracy (88.5%), sensitivity (89.6%), and specificity (86.8%).

**Table 3:** Results of binary logistic regression analyses with major depression (MDD) as dependent variables and healthy controls (HCs) as reference groups.

| Dichotomies |  | Explanatory Variables | B | SE | Wald (df=1) | p | OR | 95% CI |
| --- | --- | --- | --- | --- | --- | --- | --- | --- |
| **MDD**<br>*versus healthy*<br>**controls** | **Model #1** | Albumin | -1.897 | 0.371 | 26.170 | <0.001 | 0.150 | 0.073; 0.310 |
|  |  | ApoE | -0.606 | 0.278 | 4.739 | 0.029 | 0.545 | 0.316; 0.941 |
|  |  | C-peptide | -0.814 | 0.272 | 8.985 | 0.003 | 0.443 | 0.260; 0.754 |
|  | **Model #2** | Albumin | -1.951 | 0.460 | 17.951 | <0.001 | 0.142 | 0.058; 0.351 |
|  |  | hsCRP | -1.313 | 0.665 | 3.896 | 0.048 | 0.269 | 0.073; 0.991 |
|  |  | ApoE | -0.710 | 0.336 | 4.466 | 0.035 | 0.492 | 0.254; 0.950 |
|  |  | C-peptide | -0.776 | 0.320 | 5.904 | 0.015 | 0.460 | 0.246; 0.861 |
|  |  | ET-1 | -0.770 | 0.331 | 5.400 | 0.020 | 0.463 | 0.242; 0.886 |
|  |  | Family Dysfunction | 1.502 | 0.398 | 14.216 | <0.001 | 4.491 | 2.057; 9.807 |

Linear Discriminant Analysis (LDA) with stepwise variable selection demonstrated that five variables significantly separated the DSM diagnostic groups (Wilks’ λ = 0.447, χ^2^ = 101.047, df = 5, p < 0.001, canonical correlation = 0.744, eigenvalue = 1.237): Albumin (β= 0.581, r_s =_ 0.710), Family Dysfunction (β = -0.514, r_s =_ -0.492), C-peptide (β = 0.390, r_s =_ 0.380), Transferrin (β = 0.321, r_s =_ 0.439), and ET-1 (β= 0.317, r_s =_ 0.143). The group centroids were positioned at +1.330 (HCs) and -0.916 (MDD). The discriminant function achieved an overall classification accuracy of 83.8%, maintaining 83.1% under leave-one-out cross-validation (cross-validated sensitivity = 86.8%, specificity = 80.5%). ROC curve analysis of the predicted probabilities yielded an AUC of 0.948 (95% CI= 0.913;0.982), Gini index = 0.896, max K-S = 0.808, Youden’s index = 0.808, and an overall model quality of 0.91 at an optimal cutoff of 0.601.

### Correlations of biomarkers with demographic characteristics, early-life stressors, and clinical and behavioral phenotypes

**Figure 1** presents the heatmap of the Pearson correlation analyses. C-peptide, NO, albumin, transferrin, ApoE, and ApoA1 were negatively correlated with pure affective symptoms, the NSSI spectrum, suicidal ideation severity, physiosomatic symptoms, and self-regulation behavior scores, and positively correlated with resilience (all p < 0.05). C-peptide, NO, and albumin were also negatively correlated with family dysfunction, maladaptive controlling parenting, bullying exposure, and childhood trauma. Transferrin was negatively correlated with bullying exposure and age, ApoE with childhood trauma, and ApoA1 with family dysfunction and childhood trauma.

**Figure 1.**
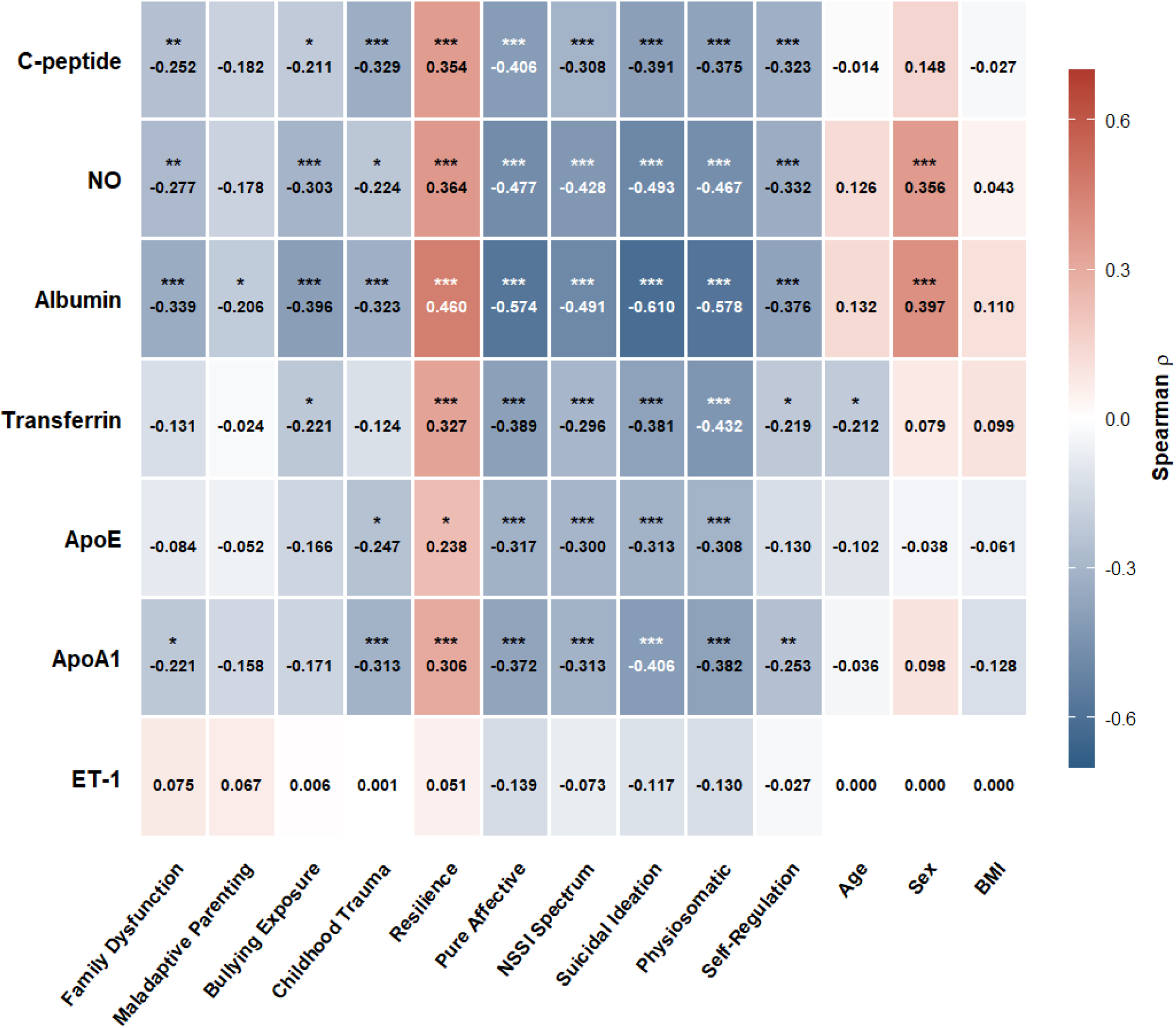
Person Correlations among biomarkers and early life in the combined sample. Cells display Person’s Correlation coefficients (*ρ*). \**p* < 0.05, \*\**p* < 0.01, ∗∗∗*p* < 0.001, two-tailed.

### Multiple regression analyses

**Table 4** presents multiple regression models for resilience and clinical and behavioral phenotypes. Model 1 explained 52.6% of the variance in resilience, with negative associations for family dysfunction and ACEs and a positive association with NAPR. Model 2 explained 55.3% of the variance in pure affective symptoms, with negative associations for albumin, C-peptide, ET-1, and ApoE and positive associations for bullying exposure and family dysfunction. Model 3 explained 46.2% of the variance in the NSSI spectrum, with a positive association for childhood trauma and a negative association for albumin. Model 4 explained 61.2% of the variance in integrated suicidal ideation severity, with negative associations for albumin and C-peptide and positive associations for childhood trauma and family dysfunction. **Figure 2 and Figure 3** shows the independent effects (partial regression) of albumin and childhood trauma, respectively, on the integrated suicidal ideation severity on after considering the effects of all other variables shown in model 4.

**Figure 2.**
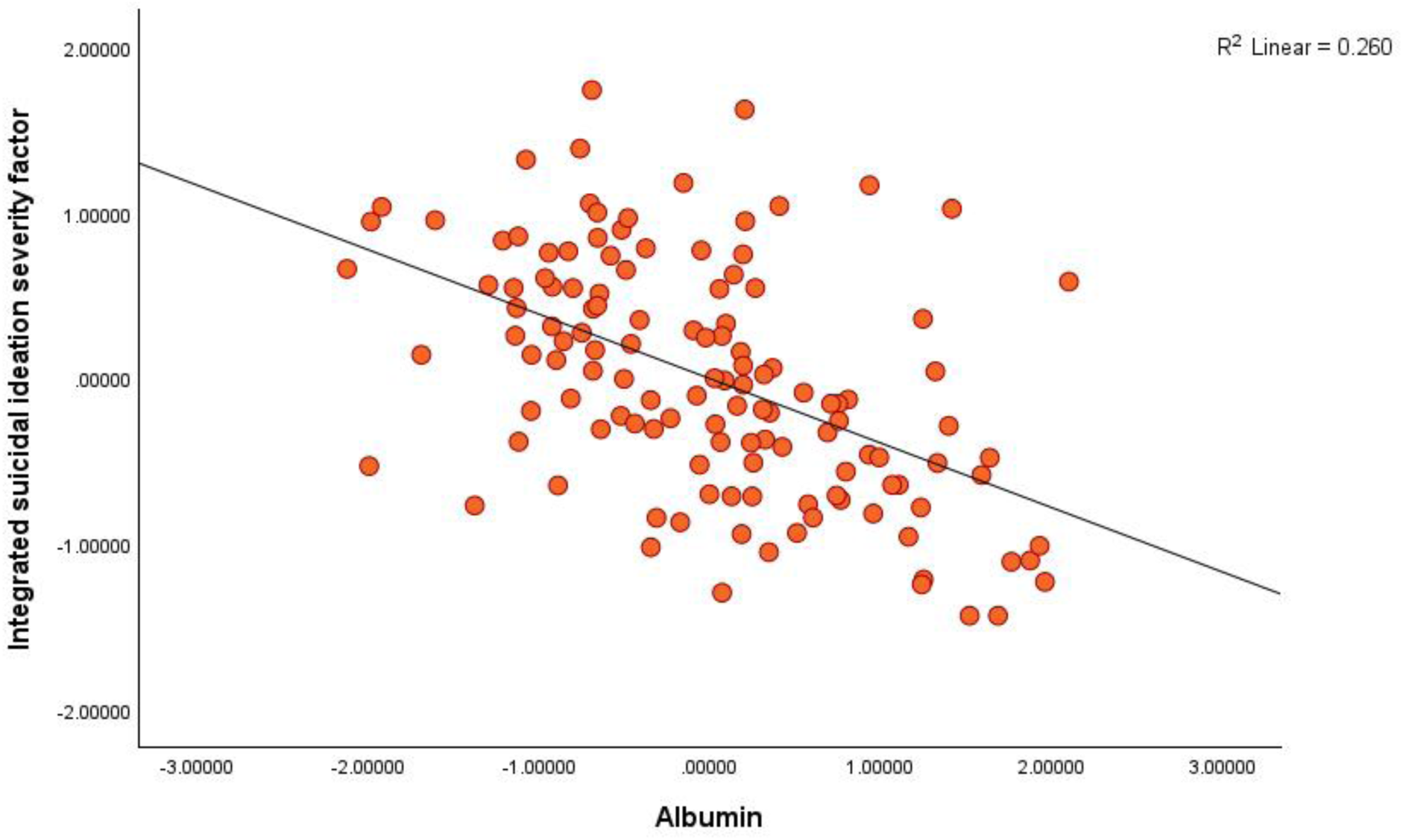
Partial regression of the Integrated Suicidal Ideation Severity on albumin levels after adjusting for childhood trauma, family dysfunction, and C-peptide (*p* < 0.001).

**Figure 3.**
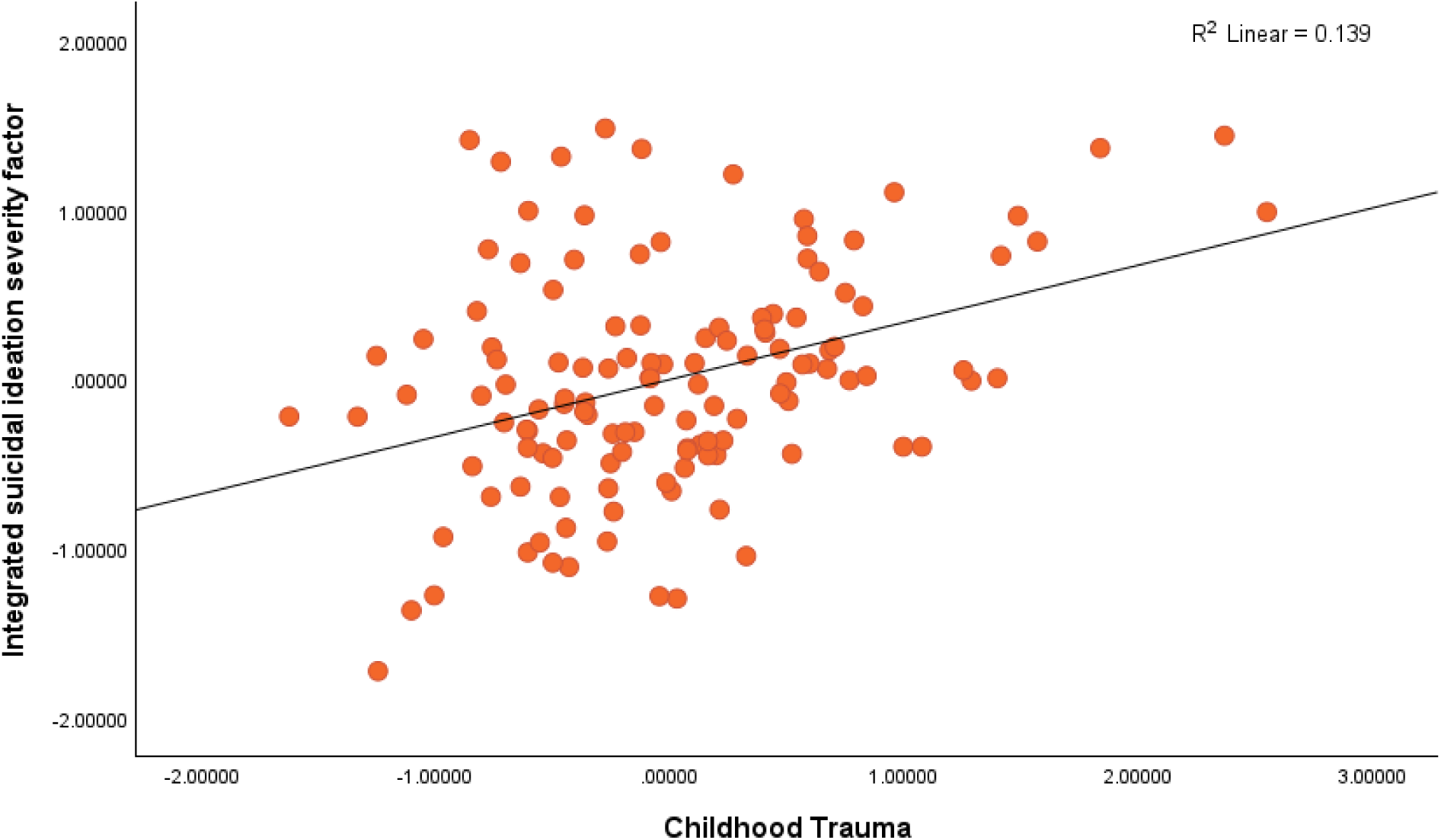
Partial regression of the Integrated Suicidal Ideation Severity on childhood trauma after adjusting for albumin, family dysfunction, and C-peptide (*p* < 0.001).

**Table 4.** Multiple regression models predict clinical and behavioural phenotypes using psychosocial adversity and biomarkers as explanatory variables.

| Dependent Variable | Predictors | Model statistics |  |  | Parameter estimates and statistics |  |  |
| --- | --- | --- | --- | --- | --- | --- | --- |
| | | R<sup>2</sup> | (df) F | p | $\beta$ | t | sig |
| #1. Resilience |  | 0.526 | (3, 125) 46.29 | <0.001 |  |  |  |
|  | Family Dysfunction |  |  |  | -0.460 | -5.836 | <0.001 |
|  | NAPR |  |  |  | 0.256 | 3.924 | <0.001 |
|  | Childhood Trauma |  |  |  | -0.196 | -2.475 | 0.015 |
| #2. Pure_affective |  | 0.553 | (6, 122) 25.13 | <0.001 |  |  |  |
|  | Albumin |  |  |  | -0.313 | -4.454 | <0.001 |
|  | C-peptide |  |  |  | -0.215 | -3.340 | 0.001 |
|  | Bullying Exposure |  |  |  | 0.204 | 2.825 | 0.006 |
|  | Family Dysfunction |  |  |  | 0.243 | 3.415 | <0.001 |
|  | ET-1 |  |  |  | -0.132 | -2.158 | 0.033 |
|  | ApoE |  |  |  | -0.128 | -1.988 | 0.049 |
| #3. NSSI Spectrum |  | 0.462 | (2, 126) 54.03 | <0.001 |  |  |  |
|  | Childhood Trauma |  |  |  | 0.502 | 7.230 | <0.001 |
|  | Albumin |  |  |  | -0.320 | -4.605 | <0.001 |
| #4. Integrated Suicidal Ideation Severity |  | 0.612 | (4, 123) 48.57 | <0.001 |  |  |  |
|  | Albumin |  |  |  | -0.402 | -6.569 | <0.001 |
|  | Childhood Trauma |  |  |  | 0.328 | 4.456 | <0.001 |
|  | Family Dysfunction |  |  |  | 0.193 | 2.669 | 0.009 |
|  | C-peptide |  |  |  | -0.138 | -2.282 | 0.024 |
| #5. Physiosomatic |  | 0.551 | (4, 124) 37.97 | <0.001 |  |  |  |
|  | Childhood Trauma |  |  |  | 0.348 | 4.497 | <0.001 |
| | | R<sup>2</sup> | (df) F | p | $\beta$ | t | sig |
|  | Albumin |  |  |  | -0.298 | -3.998 | <0.001 |
|  | Transferrin |  |  |  | -0.221 | -3.155 | 0.002 |
|  | Family Dysfunction |  |  |  | 0.164 | 2.103 | 0.037 |
| **#6. Self-Regulation Behaviour** |  | 0.537 | (4, 124) 35.88 | <0.001 |  |  |  |
|  | Bullying Exposure |  |  |  | 0.245 | 3.202 | 0.002 |
|  | Maladaptive Control Parenting |  |  |  | 0.232 | 3.041 | 0.003 |
|  | Childhood Trauma |  |  |  | 0.239 | 2.637 | 0.009 |
|  | Family Dysfunction |  |  |  | 0.199 | 2.476 | 0.015 |

Model 5 explained 55.1% of the variance in physiosomatic symptoms, with positive associations for childhood trauma and family dysfunction and negative associations for albumin and transferrin. Model 6 explained 53.7% of the variance in self-regulation behavior scores, with positive associations for bullying exposure, maladaptive controlling parenting, childhood trauma, and family dysfunction.

**Table 5** presents multiple regression models for biomarkers in relation to early-life stressors and demographic characteristics. Childhood trauma was inversely associated with C-peptide, accounting for 11.0% of its variance (model #1). Sex was positively associated, and bullying exposure negatively associated, with NO, NAPR, and albumin, with the respective models explaining 19.7%, 15.0%, and 28.4% of their variance (models #2–4). Bullying exposure and age were negatively associated with transferrin, jointly accounting for 10.5% of its variance (model #5). Childhood trauma and BMI were negatively associated with HDL-C, jointly accounting for 12.8% of its variance (model #6).

**Table 5.** Regression of biomarkers on early-life stressors and demographic covariates.

| Dependent Variable | Predictors | Model statistics |  |  | Parameter estimates and statistics |  |  |
| --- | --- | --- | --- | --- | --- | --- | --- |
| | | R<sup>2</sup> | (df) *F* | *p* | $\beta$ | *t* | *sig* |
| #1. C-Peptide |  | 0.110 | (1, 127) 15.64 | <.001 |  |  |  |
|  | Childhood Trauma |  |  |  | -0.331 | -3.955 | <0.001 |
| #2. NO |  | 0.197 | (2,126) 15.45 | <0.001 |  |  |  |
|  | Sex |  |  |  | 0.328 | 4.067 | <0.001 |
|  | Bullying Exposure |  |  |  | -0.258 | -3.203 | 0.002 |
| #3. NAPR |  | 0.150 | (2,126) 11.07 | <0.001 |  |  |  |
|  | Bullying Exposure |  |  |  | -0.305 | -3.679 | <0.001 |
|  | Sex |  |  |  | 0.200 | 2.409 | 0.017 |
| #4. Albumin |  | 0.284 | (2,126) 24.99 | <0.001 |  |  |  |
|  | Sex |  |  |  | 0.360 | 4.730 | <0.001 |
|  | Bullying Exposure |  |  |  | -0.347 | -4.563 | <0.001 |
| #5. Transferrin |  | 0.105 | (2,126) 7.41 | <0.001 |  |  |  |
|  | Bullying Exposure |  |  |  | -0.242 | -2.863 | 0.005 |
|  | Age |  |  |  | -0.236 | -2.790 | 0.006 |
| #6. HDL-C |  | 0.128 | (2,126) 9.21 | <0.001 |  |  |  |
|  | Childhood Trauma |  |  |  | -0.294 | -3.526 | <0.001 |
|  | BMI |  |  |  | -0.221 | -2.655 | 0.009 |

## Discussion

### Loss of peripheral homeostatic protection and a dissociated acute-phase response

The present study employed a systems-biology framework to examine interconnected metabolic, lipoprotein, acute-phase, oxidative, and endothelial pathways in adolescent MDD and their associations with developmental adversity and multidimensional clinical phenomes. The principal finding is a coordinated reduction in circulating albumin, transferrin, C-peptide, NO, ApoA1, ApoE, and HDL-C, accompanied by an increased ApoB/ApoA1 ratio. By contrast, hsCRP, conventional lipid indices (TG, TC, LDL-C), ApoB, AOPP, oxHDL, insulin, glucose, and ET-1 did not differ significantly between groups. These findings suggest that adolescent MDD is characterized less by overt activation of classical inflammatory, oxidative-damage, or atherogenic pathways than by a systems-level reduction in peripheral homeostatic protection, encompassing negative acute-phase proteins, metabolic regulation, lipoprotein-mediated protection, and endothelial signaling.

Diminished albumin and transferrin align with previous findings in MDD (Maes, Vandewoude et al. 1991, Almulla, Thipakorn et al. 2022, Almulla, Thipakorn et al. 2023, Almulla, Niu et al. 2025, Chen, Niu et al. 2026, Luo, Niu et al. 2026). These negative acute-phase proteins may be suppressed by chronic inflammatory signaling (Maes 1993, Khovidhunkit, Kim et al. 2004, Chojkier 2005, Chen, Niu et al. 2026). Albumin also contributes to extracellular antioxidant defense through its free thiol group and binding of redox-active molecules, whereas transferrin limits iron-dependent radical generation by sequestering ferric iron (Brieland, Clarke et al. 1992, Roche, Rondeau et al. 2008). Their coordinated reduction may therefore weaken circulating antioxidant and metal-binding defenses.

Lower albumin and transferrin without significantly increased hsCRP indicate a dissociation between the negative and positive arms of the acute-phase response. We term this profile a dissociated acute-phase response (DAPR): a pronounced reduction in negative acute-phase proteins, reflected by lower albumin, transferrin, and the NAPR composite, without a corresponding CRP-defined positive acute-phase response. Indeed, hsCRP tended to be lower in MDD and was inversely associated with MDD in multivariable models. Together with previously reported alterations in monomeric rather than pentameric CRP (Almulla, Niu et al. 2025), this pattern appears distinct from conventional CRP-dominant hepatic inflammation.

This dissociation may reflect differential cytokine regulation of negative and positive acute-phase proteins, although altered hepatic protein synthesis, amino-acid availability, nutritional status, and protein turnover may also contribute (Fuhrman, Charney et al. 2004, Gruys, Toussaint et al. 2005). The phenotype may therefore precede, or differ from, the obesity- and metabolic-syndrome-associated inflammatory phenotype characterized by elevated CRP. Within the NIMETOX framework, it is consistent with relative dissociation of immune, metabolic, and oxidative components rather than consistent uniform activation of all pathways (Maes, Almulla et al. 2025, Chen, Niu et al. 2026, Maes, Almulla et al. 2026). Although albumin and transferrin are not canonical CIRS mediators, their depletion without increased hsCRP, AOPP, or oxHDL suggests impaired protective buffering rather than prominent systemic inflammation or oxidative damage (Maes and Carvalho 2018, Maes, Vasupanrajit et al. 2025).

### Impaired lipoprotein, metabolic, and endothelial protection

The reductions in ApoA1, ApoE, and HDL-C indicate disruption of the protective lipoprotein and RCT system. ApoA1 promotes cholesterol efflux, while ApoE contributes to lipid transport and receptor-mediated lipoprotein clearance; inflammatory signaling may also suppress ApoA1 synthesis and remodel HDL composition and function (Khovidhunkit, Kim et al. 2004). Their reduction may therefore impair cholesterol efflux and RCT efficiency and weaken the anti-inflammatory, antioxidative, and endothelial-protective functions of HDL. Because ApoB was unchanged, the increased ApoB/ApoA1 ratio was driven principally by reduced ApoA1 and indicates loss of antiatherogenic protection rather than increased production of ApoB-containing particles. Unchanged triglycerides and AIP further indicate that a triglyceride-rich atherogenic phenotype was not detected, although AIP cannot exclude changes in LDL particle size, HDL function, cholesterol efflux, or LCAT activity.

These findings agree with our previous meta-analysis showing disruption of the HDL–PON1–ApoA1–LCAT/RCT system in affective disorders (Almulla, Thipakorn et al. 2023). Reduced PON1 activity may further weaken HDL protection against oxidation (Maes, Jirakran et al. 2025). Overall, the profile indicates selective loss of HDL-related and apolipoprotein protection rather than generalized dyslipidemia and suggests that the metabolic component of NIMETOX extends beyond glycemic abnormalities to impaired RCT and lipoprotein-mediated protection.

Lower C-peptide is a novel finding in adolescent MDD and agrees with previous reports in depressive illness (Yapislar, Aydogan et al. 2012, Takekawa, Kudo et al. 2019), although adult studies have reported increased or unchanged concentrations (Schweiger, Greggersen et al. 2008, Lu, Fu et al. 2014, Çakici, Bot et al. 2019). C-peptide is secreted equimolarly with insulin and provides a more direct index of endogenous insulin secretion because it largely escapes hepatic extraction (Otto-Buczkowska 2014). Lower C-peptide with unchanged insulin and glucose may suggest altered insulin secretory dynamics but does not establish β-cell failure. Its concentration depends on fasting status, concurrent glucose, recent nutrient intake, and renal clearance (Vinay, Laxmi Narsimha Rao et al. 2026), and differences in hepatic insulin extraction could contribute to the apparent discordance between C-peptide and insulin.

C-peptide is also a biologically active vascular factor. Experimental studies indicate that physiological C-peptide activates endothelial signaling, increases eNOS-dependent NO production, and reduces pro-inflammatory leukocyte–endothelial interactions, potentially through calcium-dependent eNOS activation and ERK-dependent upregulation of eNOS expression (Scalia, Coyle et al. 2000, Kitamura, Kimura et al. 2003, Forst, Hach et al. 2009). Lower C-peptide may therefore reduce metabolic–vascular support and endothelial protection.

The parallel reductions in C-peptide, ApoA1/HDL-C, and NO support a convergent disturbance in endothelial homeostatic signaling (Cheyou and Srivastava 2015, Almulla, Thipakorn et al. 2023, Dakroub, Dbouk et al. 2024). HDL promotes endothelial NO signaling through SR-BI-dependent pathways, while HDL-bound ApoM delivers sphingosine-1-phosphate to endothelial S1P receptors and supports vascular barrier integrity (Mineo and Shaul 2003, Christoffersen, Obinata et al. 2011). Thus, lower ApoA1/HDL-C and C-peptide may converge on reduced eNOS/NO support, providing a biologically plausible explanation for lower circulating NO.

Reduced NO may compromise endothelium-dependent vasodilation and endothelial anti-inflammatory, antithrombotic, antioxidant, and antiatherogenic protection (Gkaliagkousi and Ferro 2011). However, this interpretation remains inferential because eNOS activity and vascular function were not assessed in this study. Previously, Zhang et al., reported higher plasma NO levels in MDD compared to healthy controls (Lu, Zhang et al. 2018). ET-1 was unchanged univariately, although it contributed inversely to multivariable models; therefore, this is not the classical endothelial activation pattern of reduced NO with increased ET-1. Likewise, unchanged AOPP and oxHDL favor altered endothelial homeostatic signaling rather than established oxidative endothelial injury.

### Developmental adversity, peripheral dysregulation, and multidimensional MDD phenomes

The second major finding is that family dysfunction, maladaptive controlling parenting, bullying exposure, and childhood trauma were associated with lower concentrations of protective biomarkers, including C-peptide, NO, albumin, transferrin, and ApoA1. This pattern extends our previous analysis of the same cohort, in which family dysfunction and maladaptive parenting contributed to childhood trauma and multidimensional psychopathology (Wang, Wang et al. 2026). The biomarker associations were largely unrelated to BMI, whereas hsCRP was positively associated with BMI. Thus, adiposity may explain part of the variation in hsCRP (Almulla, Kitov et al. 2025, Maes, Niu et al. 2025), but does not readily explain the reductions in these protective biomarkers.

Early-life stressors and demographic factors explained 10.5%–28.4% of the variance in individual biomarkers. Childhood trauma accounted for variation in C-peptide and HDL-C, whereas bullying exposure, together with age or sex in some models, was associated with NO, albumin, transferrin, and the NAPR composite. Different forms of adversity may therefore have partly distinct biological correlates, although the cross-sectional design cannot establish temporal order, biological programming, or stressor-specific mechanisms.

The protective biomarkers were inversely associated with pure affective symptoms, the NSSI spectrum, suicidal ideation severity, physiosomatic symptoms, and self-regulation behavior scores, and positively associated with resilience. Models combining early-life adversity with biomarkers explained 46.2%–61.2% of the variance in resilience and the clinical and behavioral phenomes. Albumin, transferrin, C-peptide, ApoE, and ET-1 retained associations with specific phenomes after accounting for relevant adversity measures. Childhood trauma was particularly associated with the NSSI spectrum, whereas childhood trauma and family dysfunction were associated with suicidal ideation and physiosomatic symptoms. In contrast, the self-regulation behavior model was explained by psychosocial stressors alone. This domain specificity argues against treating MDD as a single binary endpoint and suggests that distinct biological dimensions map onto different clinical phenomes.

These findings agree with studies linking adverse childhood experiences, negative acute-phase proteins, lipid disturbances, and NIMETOX pathways to affective, suicidal, and physiosomatic phenotypes (Maes, Rachayon et al. 2022, Almulla, Niu et al. 2025, Almulla, Niu et al. 2026, Chen, Niu et al. 2026, Luo, Niu et al. 2026). Other studies show that adverse childhood experiences and negative life events are associated with sensitized immune-inflammatory and NIMETOX networks, diminished compensatory immunoregulation, and disturbed lipid metabolism, which statistically explained part of the association between adversity and multidimensional affective phenotypes (Maes, Rachayon et al. 2022, Almulla, Algon et al. 2024, Sirivatanapa, Janta et al. 2026). These observations support interacting immune, metabolic, and lipid pathways, but do not demonstrate mediation in the present study.

### Integrated NIMETOX interpretation

The findings support an integrated model in which developmental adversity-associated allostatic dysregulation is linked to reduced systemic homeostatic reserve. Family dysfunction, childhood trauma, bullying, and maladaptive parenting were associated with disturbances across several protective domains: lower albumin and transferrin; reduced ApoA1, ApoE, and HDL-C with a higher ApoB/ApoA1 ratio; lower C-peptide; and reduced NO without increased ET-1. Collectively, these alterations may define a NIMETOX homeostatic protection deficit.

The present data support associations among these domains, not the proposed causal sequence. We therefore propose that adversity-associated allostatic dysregulation may contribute to loss of systemic homeostatic reserve, thereby reducing biological and psychological resilience and increasing vulnerability to affective, physiosomatic, NSSI, suicidal, and self-regulation phenomes. Adolescent MDD thus may be characterized less by excess activation of damaging NIMETOX pathways than by deficient protective and homeostatic systems. We hypothesize that this adolescent homeostatic protection deficit may constitute an early vulnerability phenotype that, in susceptible individuals, precedes recurrent depressive episodes and the more pronounced NIMETOX abnormalities associated with recurrence of illness (ROI) in adult MDD (Maes, Almulla et al. 2025). This proposed developmental trajectory remains to be established in prospective studies.

### Classification performance of integrated nomothetic models

The third major finding is that multivariable models combining selected metabolic, vascular, and negative acute-phase biomarkers, with or without developmental adversity measures, achieved classification accuracies of 82.6%–88.5%. Lower albumin, C-peptide, and ApoE were particularly informative in the biomarker-only logistic model, whereas the inclusion of hsCRP (inversely associated), ET-1 (inversely associated), and family dysfunction (positively associated) improved classification performance in the extended model.

This performance is consistent with previous nomothetic models combining inflammatory, acute-phase, lipid, and psychological adversity measures to discriminate MDD from healthy controls (Almulla, Niu et al. 2025, Almulla, Niu et al. 2026, Chen, Niu et al. 2026). The findings indicate that adolescent MDD may be characterized more accurately by combining metabolic, vascular, lipid, and developmental information than by relying on a single biomarker domain. Notably, hsCRP was the least significant factor in model 2, consistent with previous research excluding hsCRP from the MDD biomarker panel (Maes, Niu et al. 2025).

These models should be considered internally validated classification models, not diagnostic tests. Clinical utility requires external validation, calibration analysis, comparison with clinical-only models, and testing in psychiatric control groups and clinically defined subgroups.

## Limitations

The findings should be interpreted in light of several limitations. First, the cross-sectional observational design precludes causal inference. Longitudinal cohorts are required to determine whether peripheral immunometabolic and vascular changes precede the progression of affective illness or emerge as a consequence of it. Second, the cohort comprised acutely hospitalized Chinese adolescents without overt cardiometabolic comorbidity. This design identifies a distinct biological profile in youth, but the findings may not generalize to older or chronically ill patients, individuals with metabolic syndrome, or community and outpatient adolescent samples. Third, although key covariates such as age, sex, and BMI were considered, nutritional, hepatic, renal, medication-related, and environmental influences could not be fully accounted for. Fourth, direct measures of vascular function, including eNOS phosphorylation, endothelial glycocalyx shedding markers, and flow-mediated dilation, were not available. Consequently, the endothelial interpretation remains inferential. Finally, future studies should assess LCAT and PON1 activity to assess the proposed impairment in RCT and HDL-related antioxidant protection.

## Conclusion

Adolescent MDD was characterized by a coordinated reduction in C-peptide, NO, negative acute-phase proteins, ApoA1, ApoE, and HDL-C, together with an increased ApoB/ApoA1 ratio, without classical CRP elevation or a metabolic-syndrome lipid profile. This pattern suggests a deficit in peripheral metabolic, antioxidant, lipoprotein, and endothelial homeostatic protection. Its associations with early developmental adversity, resilience, and specific clinical phenomes support an integrated NIMETOX model in which reduced protective reserve contributes to vulnerability across affective, physiosomatic, suicidal, NSSI, and self-regulation dimensions. Integrated biological and psychosocial models also showed very good internal accuracy. Longitudinal and externally validated studies are needed to determine temporal direction, clinical specificity, and whether this early protective deficit precedes the more overt NIMETOX damage phenotype observed in recurrent adult MDD.

## Authors’ contributions

Conceptualization and study design: Michael Maes and Abbas F. Almulla; first draft writing: Abbas F. Almulla; biomarker measurement: Huajie Mao, Abbas F. Almulla, and Yingqian Zhang; statistical analysis: Michael Maes; editing: all authors; recruitment of patients: Ping Wang, Yaoyin Zhang, Juan Li, Junmin Chen, and Yulan Huang. All authors approved the final version of the manuscript.

## Ethics approval and consent to participate

The study protocol was approved by the Ethics Committee of the University of Electronic Science and Technology of China (Approval No. 31682). All participants aged 18 years provided written informed consent. For participants younger than 18 years, written informed consent was obtained from a parent or legal guardian, along with written assent from the adolescent.

## Data Availability Statement

The datasets of the current study will be available from the corresponding author upon reasonable request once the study’s primary research aims have been completed by the authors.

## References

Almulla, A. F., A. A. A. Algon and M. Maes (2024). “Adverse childhood experiences and recent negative events are associated with activated immune and growth factor pathways, the phenome of first episode major depression and suicidal behaviors.” Psychiatry Research 334: 115812.

Almulla, A. F., S. Kitov, T. Deneva, M. F. Kitova, L. Kitova, K. Stoyanova, D. Stoyanov and M. Maes (2025). “C-reactive protein is not a biomarker of depression severity in drug-naïve obese patients with metabolic syndrome.” Acta Neuropsychiatr: 1–43.

Almulla, A. F. and M. Maes (2025). “Peripheral Immune-Inflammatory Pathways in Major Depressive Disorder, Bipolar Disorder, and Schizophrenia: Exploring Their Potential as Treatment Targets.” CNS Drugs 39(8): 739–762.

Almulla, A. F., M. Niu, Y. Luo, T. Chen, C. Yangyang, Y. Zhang and M. Maes (2026). “Increased oxidation susceptibility of HDL particles as a mechanistic signature of major depressive disorder.” Advances in Redox Research 19: 100163.

Almulla, A. F., M. Niu, D. Stoyanov, Y. Zhang and M. Maes (2025). “Monomeric CRP and negative acute phase proteins, but not pentameric CRP, as biomarkers of major depression and MDMD.” Acta Neuropsychiatrica: 1–30.

Almulla, A. F., Y. Thipakorn, A. A. A. Algon, C. Tunvirachaisakul, H. K. Al-Hakeim and M. Maes (2023). “Reverse cholesterol transport and lipid peroxidation biomarkers in major depression and bipolar disorder: A systematic review and meta-analysis.” Brain Behav Immun 113: 374–388.

Almulla, A. F., Y. Thipakorn, A. Vasupanrajit, A. A. Abo Algon, C. Tunvirachaisakul, A. A. Hashim Aljanabi, G. Oxenkrug, H. K. Al-Hakeim and M. Maes (2022). ”The tryptophan catabolite or kynurenine pathway in major depressive and bipolar disorder: A systematic review and meta-analysis.” Brain, Behavior, & Immunity - Health 26: 100537.

Beck, A. T., M. Kovacs and A. Weissman (1979). “Assessment of suicidal intention: the Scale for Suicide Ideation.” Journal of consulting and clinical psychology 47(2): 343.

Bernstein, D. P., J. A. Stein, M. D. Newcomb, E. Walker, D. Pogge, T. Ahluvalia, J. Stokes, L. Handelsman, M. Medrano and D. Desmond (2003). “Development and validation of a brief screening version of the Childhood Trauma Questionnaire.” Child abuse & neglect 27(2): 169–190.

Birmaher, M. B., M. S. Khetarpal, M. D. Brent, M. M. Cully, M. L. Balach, P. D. J. Kaufman And P. D. S. M. Neer (1997). “The screen for child anxiety related emotional disorders (SCARED): Scale construction and psychometric characteristics.” Journal of the American academy of child & adolescent psychiatry 36(4): 545–553.

Brieland, J. K., S. J. Clarke, S. Karmiol, S. H. Phan and J. C. Fantone (1992). “Transferrin: A potential source of iron for oxygen free radical-mediated endothelial cell injury.” Archives of Biochemistry and Biophysics 294(1): 265–270.

Cabou, C., P. Honorato, T. Duparc, A. Fournel, L. Briceño, M. León, L. Frayssinhe, N. Parada, V. Aguilera, C. Aguayo, C. Knauf, M. González, L. Martinez and C. Radojkovic (2017). “Apolipoprotein A-I increases nitric oxide synthesis and vasodilator response in vivo through the activation of Ecto-F1-ATPase / P2Y1 axis.” Atherosclerosis 263: e6.

Çakici, N., M. Bot, F. Lamers, T. Janssen, P. J. van der Spek, L. de Haan, S. Bahn, B. W. J. H. Penninx and N. J. M. van Beveren (2019). “Increased serum levels of leptin and insulin in both schizophrenia and major depressive disorder: A cross-disorder proteomics analysis.” European Neuropsychopharmacology 29(7): 835–846.

Cellini, M., E. Strobbe, C. Gizzi, N. Balducci, P. G. Toschi and E. C. Campos (2012). “Endothelin-1 plasma levels and vascular endothelial dysfunction in primary open angle glaucoma.” Life Sciences 91(13): 699–702.

Chen, T., M. Niu, Y. Luo, J. Li, A. F. Almulla, Y. Zhang and M. Maes (2025). “The acute phase inflammatory response as a key determinant of reduced lipid-associated antioxidant defenses in Chinese patients with major depressive disorder.” medRxiv: 2025.2010.2008.25337557.

Chen, T., M. Niu, Y. Luo, J. Li, A. F. Almulla, Y. Zhang and M. Maes (2026). “The acute phase inflammatory response as a key determinant of reduced lipid-associated antioxidant defenses in Chinese patients with major depressive disorder.” Brain, Behavior, and Immunity 138: 106923.

Cheyou, E. R. S. and A. K. Srivastava (2015). Vascular Endothelial Dysfunction and Atherosclerosis: Role of Nitric Oxide System. <u>Pathophysiology and Pharmacotherapy of Cardiovascular Disease</u>. G. Jagadeesh, P. Balakumar and K. Maung-U. Cham, Springer International Publishing: 547–561.

Chojkier, M. (2005). “Inhibition of albumin synthesis in chronic diseases: molecular mechanisms.” J Clin Gastroenterol 39(4 Suppl 2): S143–146.

Christoffersen, C., H. Obinata, S. B. Kumaraswamy, S. Galvani, J. Ahnström, M. Sevvana, C. Egerer-Sieber, Y. A. Muller, T. Hla, L. B. Nielsen and B. Dahlbäck (2011). “Endothelium-protective sphingosine-1-phosphate provided by HDL-associated apolipoprotein M.” Proc Natl Acad Sci U S A 108(23): 9613–9618.

Connor, K. M. and J. R. Davidson (2003). “Development of a new resilience scale: The Connor-Davidson resilience scale (CD-RISC).” Depression and anxiety 18(2): 76–82.

Dakroub, A., A. Dbouk, A. Asfour, S. A. Nasser, A. F. El-Yazbi, A. Sahebkar, A. A. Eid, R. Iratni and A. H. Eid (2024). “C-peptide in diabetes: A player in a dual hormone disorder?” J Cell Physiol 239(5): e31212.

Epstein, N. B., L. M. Baldwin and D. S. Bishop (1983). “The McMaster family assessment device.” Journal of marital and family therapy 9(2): 171–180.

Forst, T., T. Hach, T. Kunt, M. M. Weber and A. Pfützner (2009). “Molecular effects of C-Peptide in microvascular blood flow regulation.” Rev Diabet Stud 6(3): 159–167.

Fuhrman, M. P., P. Charney and C. M. Mueller (2004). “Hepatic proteins and nutrition assessment.” Journal of the American Dietetic Association 104(8): 1258–1264.

Gkaliagkousi, E. and A. Ferro (2011). “Nitric oxide signalling in the regulation of cardiovascular and platelet function.” Front Biosci (Landmark Ed) 16(5): 1873–1897.

Gruys, E., M. J. Toussaint, T. A. Niewold and S. J. Koopmans (2005). “Acute phase reaction and acute phase proteins.” J Zhejiang Univ Sci B 6(11): 1045–1056.

Hamilton, M. (1959). “The assessment of anxiety states by rating.” British journal of medical psychology.

Hamilton, M. (1960). “A rating scale for depression.” Journal of neurology, neurosurgery, and psychiatry 23(1): 56.

Khovidhunkit, W., M. S. Kim, R. A. Memon, J. K. Shigenaga, A. H. Moser, K. R. Feingold and C. Grunfeld (2004). “Effects of infection and inflammation on lipid and lipoprotein metabolism: mechanisms and consequences to the host.” J Lipid Res 45(7): 1169–1196.

Kitamura, T., K. Kimura, K. Makondo, D. T. Furuya, M. Suzuki, T. Yoshida and M. Saito (2003). “Proinsulin C-peptide increases nitric oxide production by enhancing mitogen-activated protein-kinase-dependent transcription of endothelial nitric oxide synthase in aortic endothelial cells of Wistar rats.” Diabetologia 46(12): 1698–1705.

Klein, D. N., B. A. Arnow, J. L. Barkin, F. Dowling, J. H. Kocsis, A. C. Leon, R. Manber, B. O. Rothbaum, M. H. Trivedi and S. R. Wisniewski (2009). “Early adversity in chronic depression: clinical correlates and response to pharmacotherapy.” Depress Anxiety 26(8): 701–710.

Kovacs, M. (1992). “Children’s depression inventory.” Acta Paedopsychiatrica: International Journal of Child & Adolescent Psychiatry.

Lu, Y. R., X. Y. Fu, L. G. Shi, Y. Jiang, J. L. Wu, X. J. Weng, Z. P. Wang, X. Y. Wu, Z. Lin, W. B. Liu, H. C. Li, J. H. Luo and A. M. Bao (2014). “Decreased plasma neuroactive amino acids and increased nitric oxide levels in melancholic major depressive disorder.” BMC Psychiatry 14: 123.

Luo, Y., M. Niu, T. Chen, J. Li, A. F. Almulla, Y. Zhang and M. Maes (2026). “Lower insulin resistance in Chinese patients with severe major depressive disorder: associations with the inflammatory response.” BMC Psychiatry 26(1).

Maes, M. (1993). “A review on the acute phase response in major depression.” Rev Neurosci 4(4): 407–416.

Maes, M., A. F. Almulla, D. Stoyanov and Y. Zhang (2025). “Advancements in the molecular understanding of major depressive disorder uncovering novel targets with therapeutic promise: focus on recurrence of illness.” Expert Opin Ther Targets 29(11-12): 775–791.

Maes, M., A. F. Almulla, D. Stoyanov and Y. Zhang (2026). “Unifying the hallmarks of major depression through neuroimmune–metabolic–oxidative (NIMETOX) dysregulation: a mechanistic systems framework.” Cellular & Molecular Immunology.

Maes, M., A. F. Almulla, Z. You and Y. Zhang (2025). “Neuroimmune, metabolic and oxidative stress pathways in major depressive disorder.” Nature Reviews Neurology 21(9): 473–489.

Maes, M. and A. F. Carvalho (2018). “The Compensatory Immune-Regulatory Reflex System (CIRS) in Depression and Bipolar Disorder.” Molecular Neurobiology 55(12): 8885–8903.

Maes, M., K. Jirakran, L. O. Semeão, A. P. Michelin, A. K. Matsumoto, F. F. Brinholi, D. S. Barbosa, C. Tivirachaisakul, A. F. Almulla, D. Stoyanov and Y. Zhang (2025). “Key factors underpinning neuroimmune-metabolic-oxidative (NIMETOX) major depression in outpatients: paraoxonase 1 activity, reverse cholesterol transport, increased atherogenicity, protein oxidation, and differently expressed cytokine networks.” Neuro Endocrinol Lett 46(2): 115–125.

Maes, M., M. Niu, X. Zhang, J. Li, D. Stoyanov, B. Zhou, A. F. Almulla and Y. Zhang (2025). “The negative acute phase response and not serum C-reactive protein is a major biomarker of major depression: a precision nomothetic psychiatry study.” medRxiv: 2025.2008.2015.25333787.

Maes, M., M. Rachayon, K. Jirakran, P. Sodsai, S. Klinchanhom, M. Debnath, A. Basta-Kaim, M. Kubera, A. F. Almulla and A. Sughondhabirom (2022). “Adverse Childhood Experiences Predict the Phenome of Affective Disorders and These Effects Are Mediated by Staging, Neuroimmunotoxic and Growth Factor Profiles.” Cells 11(9): 1564.

Maes, M., M. Vandewoude, S. Scharpé, L. De Clercq, W. Stevens, L. Lepoutre and C. Schotte (1991). “Anthropometric and biochemical assessment of the nutritional state in depression: evidence for lower visceral protein plasma levels in depression.” J Affect Disord 23(1): 25–33.

Maes, M., A. Vasupanrajit, K. Jirakran, B. Zhou, C. Tunvirachaisakul and A. F. Almulla (2025). “Simple dysmood disorder, a mild subtype of major depression, is not an inflammatory condition: Depletion of the compensatory immunoregulatory system.” Journal of Affective Disorders 375: 75–85.

Mineo, C. and P. W. Shaul (2003). “HDL stimulation of endothelial nitric oxide synthase: a novel mechanism of HDL action.” Trends Cardiovasc Med 13(6): 226–231.

Niu, M., Y. Zhang, X. Zhang, C. Yangyang, X. Zhuang, J. Li and M. Maes (2025). “Dissection of the clinical phenome of major depressive disorder into subdomains and subgroups in relation to activated immune-inflammatory profiles.” medRxiv: 2025.2010.2021.25338439.

Otto-Buczkowska, E. (2014). ”The clinical utility of C-peptide measurement in diabetology.” Pediatric Endocrinology, Diabetes and Metabolism 20(2): 63–68.

Posner, K., G. K. Brown, B. Stanley, D. A. Brent, K. V. Yershova, M. A. Oquendo, G. W. Currier, G. A. Melvin, L. Greenhill, S. Shen and J. J. Mann (2011). “The Columbia-Suicide Severity Rating Scale: initial validity and internal consistency findings from three multisite studies with adolescents and adults.” Am J Psychiatry 168(12): 1266–1277.

Roche, M., P. Rondeau, N. R. Singh, E. Tarnus and E. Bourdon (2008). “The antioxidant properties of serum albumin.” FEBS Lett 582(13): 1783–1787.

Scalia, R., K. M. Coyle, B. J. Levine, G. Booth and A. M. Lefer (2000). “C-peptide inhibits leukocyte-endothelium interaction in the microcirculation during acute endothelial dysfunction.” The FASEB Journal 14(14): 2357–2364.

Schweiger, U., W. Greggersen, S. Rudolf, M. Pusch, T. Menzel, S. Winn, J. Hassfurth, E. Fassbinder, K. G. Kahl, K. M. Oltmanns, F. Hohagen and A. Peters (2008). “Disturbed glucose disposal in patients with major depression; application of the glucose clamp technique.” Psychosom Med 70(2): 170–176.

Sirivatanapa, V., P. Janta, A. F. Almulla, A. Vasupanrajit, C. Tunvirachaisakul, S. Sriswasdi, R. Tansawat, A. F. Carvalho, Y. Zhang and M. Maes (2026). “Psychological stress-associated ceramide and diacylglyceride lipotoxicity as contributors to first-episode depression pathophysiology: A neuroimmune-metabolic-oxidative stress (NIMETOX) perspective.” J Affect Disord 414: 122351.

Swanson, J. M. (1992). School-based assessments and interventions for ADD students, KC publishing.

Takekawa, D., T. Kudo, J. Saito, F. Kimura, Y. Nikaido, K. Sawada, N. Yasui-Furukori and K. Hirota (2019). “Higher plasma leptin and lower C-peptide levels are associated with depression: A cross-sectional study.” Journal of Affective Disorders 243: 70–74.

Tonhajzerova, I., Z. Visnovcova, I. Ondrejka, D. Funakova, I. Hrtanek and N. Ferencova (2022). “Major depressive disorder at adolescent age is associated with impaired cardiovascular autonomic regulation and vasculature functioning.” Int J Psychophysiol 181: 14–22.

Vinay, E. S., B. Laxmi Narsimha Rao, N. Saf and D. Gautam (2026). “C-peptide in Precision Diabetes Care and Beyond: A Comprehensive Review.” Clinical Medicine Insights: Endocrinology and Diabetes 19: 11795514251397811.

Wallerath, T., T. Kunt, T. Forst, E. I. Closs, R. Lehmann, T. Flohr, M. Gabriel, D. Schäfer, A. Göpfert, A. Pfützner, J. Beyer and U. Förstermann (2003). “Stimulation of endothelial nitric oxide synthase by proinsulin C-peptide.” Nitric Oxide 9(2): 95–102.

Wang, P., P. Wang, Y. Zhang, X. Wang, C. Li, Y. Huang and M. Maes (2026). “Family-related experiences, childhood trauma, and cognitive-emotional correlates across psychopathological domains in adolescent depression.” medRxiv: 2026.2007.2027.26358988.

Wang, X., L. Mendelsohn, L. Freeman, B. Vaisman, A. Remaley and G. J. Kato (2014). “Stimulation of Nitric Oxide Synthase Activity By Plasma Apolipoproteins: a Biomarker of Endothelial Function in Adults with Sickle Cell Disease.” Blood 124(21): 4015–4015.

Yapislar, H., S. Aydogan and Ü. Ozüm (2012). “Biological understanding of the cardiovascular risk associated with major depression and panic disorder is important.” International Journal of Psychiatry in Clinical Practice 16(1): 27–32.

Zhou, B. and Z. Fang (1993). “Parent-child relationship diagnostic test (pcrt),.” Psychological assessment instrument, East China Normal University.

